# Reduced turnaround times through multi-sectoral collaboration during the first surge of SARS-CoV-2 in Louisiana, March-April 2020

**DOI:** 10.1101/2020.07.17.20156158

**Authors:** RC Christofferson, Hollis R. O’Neal, Tonya Jagneaux, Catherine O’Neal, Christine S. Walsh, E. Handly Mayton, Luan D. Vu, Abigail I. Fish, Anh Phan, Thaya E. Stoufflet, Jonathan R. Schroeder, Morgan Walker, Erik A. Turner, Christi Pierce, K. Scott Wester, Connie DeLeo, Edgardo Tenreiro, Beverly Ogden, Stephania A. Cormier

## Abstract

**Background:** In March 2020, an influx of admissions in COVID-19 positive patients threatened to overwhelm healthcare facilities in East Baton Rouge Parish, Louisiana. Exacerbating this problem was a shortage of diagnostic testing capability, resulting in a delay in time-to-result return. An improvement in diagnostic testing availability and timeliness was necessary to improve the allocation of resources and ultimate throughput of patients. The management of a COVID-19 positive patient or patient under investigation requires infection control measures that can quickly consume personal protective equipment (PPE) stores and personnel available to treat these patients. Critical shortages of both PPE and personnel also negatively impact care in patients admitted with non-COVID-19 illnesses.

**Methods:** A multisectoral partnership of healthcare providers, facilities and academicians created a molecular diagnostic lab within an academic research facility dedicated to testing inpatients and healthcare personnel for SARS-CoV-2. The purpose of the laboratory was to provide a temporary solution to the East Baton Rouge Parish healthcare community until individual facilities were self-sustaining in testing capabilities. We describe the partnership and the impacts of this endeavor by developing a model derived from a combination of data sources, including electronic health records, hospital operations, and state and local resources.

**Findings:** Our model demonstrates two important principles: the impact of reduced turnaround times (TAT) on potential differences in inpatient population numbers for COVID-19 and savings in PPE attributed to the more rapid TAT.

**Interpretation:** Overall, we provide rationale for and demonstration of the utility of multisectoral partnerships when responding to public health emergencies.

## Introduction

SARS-CoV-2, the etiologic agent of the disease known as COVID-19, is a member of the *Betacoronavirus* genus in the *Coronaviridae* family^1-3^. Following the first reported case of COVID-19 in December of 2019 in Wuhan, China, the virus spread rapidly globally. On January 19, 2020 the first presumptive COVID-19 case in the United States occurred in the state of Washington, ^4^ and, on March 17, 2020, East Baton Rouge Parish (EBRP) announced its first case of COVID-19.

Outbreaks place extraordinary stress on healthcare systems, and a primary goal of public health response is to avoid accelerated utilization and depletion of resources ^5,6^. Data from the Italian outbreak demonstrated that treatment of COVID-19 rapidly exhausted such resources as ICU beds, ventilators, and personnel ^7-9^. The short period of time in which these cases presented to healthcare facilities inhibited the ability to expand hospital capacity and simultaneously consumed materials through thinly stretched supply chains ^7-9^. The management of a COVID-19 positive patient or Patient Under Investigation (PUI) requires infection control measures including strict PPE utilization and, and, due to the complexity of patient care, under ideal labor supply conditions, a reduced patient to nursing staff ratio. Early depletion of resources due to an influx of patients may result in rationing of needed interventions and, ultimately, consideration of crisis standards of care. By late-March 2020, an influx of COVID-19 positive patients threatened to overwhelm the two major hospitals in COVID-19-unit, which represent 75% of the healthcare market’s inpatient admissions.

In both of these hospitals, clinicians admitted PUI’s to cohorted units until confirmation of by SARS-CoV-2 nucleic acid amplification. Because the hospitals maintained PUI’s in COVID-specific units, and because there was no specific treatment for COVID-19, a positive result for SARS-CoV-2 did not impact individual patient care; however, clinicians acted on the presumptive negative results with admission to a non-COVID-19-unit. This action often resulted in unencumbered pursual of the differential diagnosis, relief of PPE use, and bed turnover for subsequent COVID-19 patients ^10,11^.

During the first weeks of the outbreak, there was a shortage of diagnostic testing ability in the EBRP area. The Louisiana Office of Public Health Laboratory became overwhelmed with specimens from across the state, and commercial laboratories were overwhelmed with specimens from around the country. Furthermore, the local hospitals did not have in-house capability for SARS-CoV-2 testing. However, the Louisiana State University School of Veterinary Medicine (LSU SVM), housed faculty with expertise in molecular diagnostics, including real-time Reverse Transcription Polymerase Chain Reaction (RT-PCR) and human respiratory pathobiology and viral pathogens. Recognizing the unique capabilities of these faculty and their laboratory, in an effort to improve local diagnostic testing capacity and turnaround time (TAT), a multisectoral partnership of healthcare providers, facilities, and academicians repurposed academic research facilities as a dedicated COVID-19 inpatient clinical testing entity, named River Road Testing Lab (RRTL).

The following analysis of the RRTL demonstrates how research laboratories can complement existing diagnostic infrastructure in times of crisis due to an emerging infection/ pandemic, until commercial and hospital-based laboratories are able to bear the burden of testing. Further, a simple data-driven model demonstrates the differences in COVID-19 inpatient population numbers and PPE usage in scenarios where a RRTL-like, dedicated laboratory with accelerated TAT compared to the scenario where no RRTL-like laboratory exists. The study of the impact of the RRTL on the Baton Rouge area was approved by the LSU Health Sciences Center – New Orleans Institutional Review Board (LSU HSC IRB #20-043).

## Materials and Methods

### RRTL Formation

The creation of RRTL began in early March 2020 (Figure 1), prior to the first confirmed case in the COVID-19-units, by exploring supply chain options for testing reagents and supplies. On March 16, physician-scientists from the area hospitals approached the LSU SVM faculty about both supplying viral transport media (VTM) and the potential for diagnostic testing. Also, on this date, the FDA issued a “guidance to provide a policy to help accelerate the availability of novel coronavirus (COVID-19) diagnostic tests developed by laboratories and commercial manufacturers during the public health emergency ^12^.” With administrative assistance from area hospitals for navigation of the federal regulations and certification processes, RRTL received the required regulatory approvals including CLIA certification and an Emergency Use Authorization (EUA) Protocol for a lab developed test was submitted to the U.S. Food and Drug Administration. These efforts allowed for RRTL to begin testing clinical samples on March 23, 2020.

**Figure 1:**
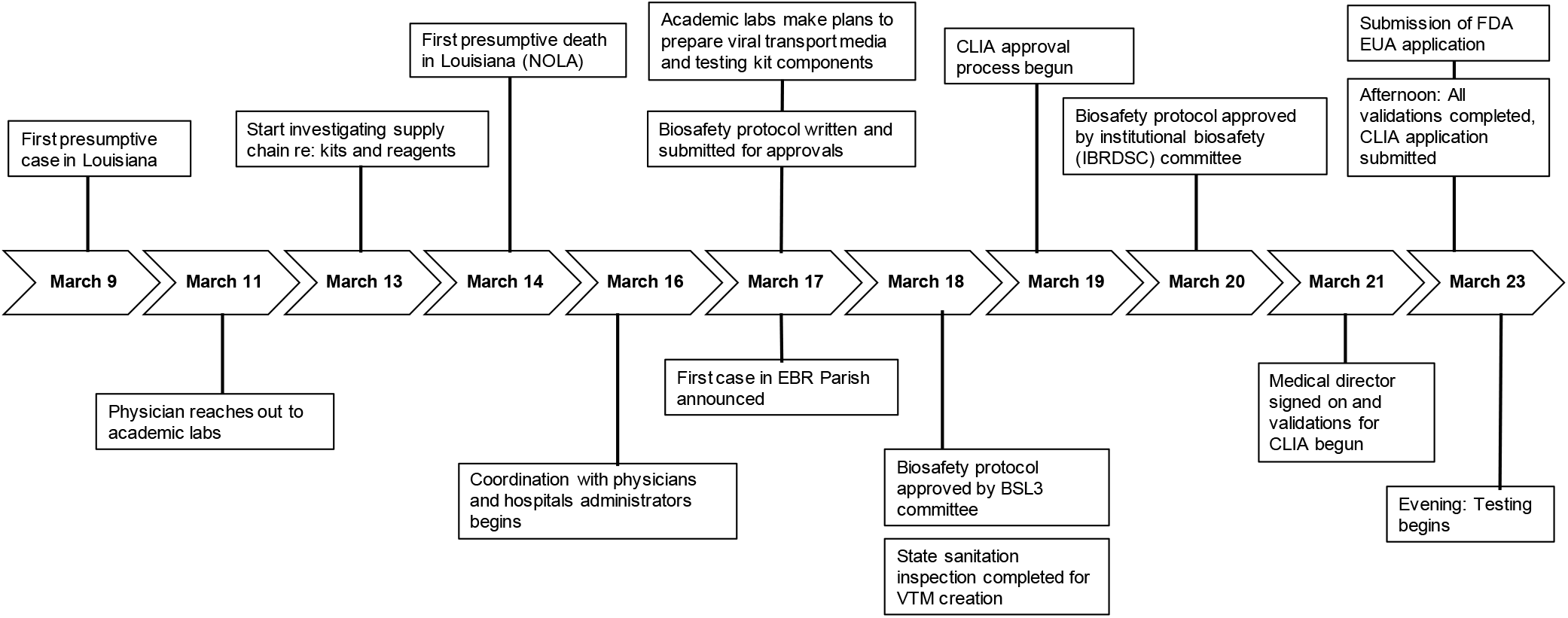
Timeline for stand-up of River Road Testing Laboratory (RRTL) at Louisiana State University.

RRTL served two major purposes during this first wave of COVID-19 in EBRP: supply of viral testing kits and testing of inpatients. First, RRTL made and distributed nearly 8,000 viral testing kits consisting of tubes, swabs, and VTM to local healthcare, with assistance from volunteers from hospital and other academic partners. These kits were critically important, as they enabled testing despite compromised supply chains amid the growing epidemic in Europe, Washington State, New York City (NYC), and elsewhere ^13,14^. RRTL performed the majority of its assays on these testing kits. RRTL received strong support from LSU administration as well as state and federal representatives of Louisiana. The Baton Rouge Area Foundation, a local area philanthropic organization, Our Lady of the Lake Regional Medical Center, and Baton Rouge General Medical Center provided financial and personnel support.

The day-to-day testing team operated under the direction of a clinical laboratory medical director. The testing team was comprised of two senior scientists, two post-doctoral researchers, three research associates, and three graduate students. The clinical team consisted of three faculty physicians and one chief resident. In addition, data and patient information were collected and managed using REDCap electronic data capture tools hosted at LSUHSC School of Public Health – New Orleans, with data entry performed by volunteers and team members from the hospitals and clinical partners ^15,16^. The process flow of samples tested by RRTL on a typical day is shown in Figure S1.

The typical daily testing capacity was 95 unique samples, with a maximum capacity of 190 samples. This meant that RRTL also was able to run samples from additional healthcare facilities, institutions, as well as healthcare workers and first responders. RRTL was the primary testing site for the area’s two largest hospitals from March 23 through April 22, 2020. The period between April 23 and May 15 served as a transition period, when local hospitals were increasing in-house testing capacity. By May 15, RRTL had run 3,857 samples since opening on March 23. The overall positivity rate was 33%, including 23.6% of hospitalized inpatients, meaning that RRTL was able to quickly provide operationally actionable results to move over 76.4% of tested patients out of COVID-19 inpatient units, provided there was no clinical or operational indication to keep the patient in the unit. The ages ranged from a premature infant to a 99-year-old patient. The TAT (defined as the time of collection to the time of result reporting) for the period of March 23, 2020 to May 15, 2020 was median of 1.67 days from the time of collection (IQR [1.6, 2.63]).

### Data and Model formulation

To estimate the impact of faster TAT due to the formation of RRTL, a model was developed to assess the time of March 23 – April 22, 2020 when RRTL was the primary testing laboratory for two major hospitals. Daily admissions of PUI’s are from a hospital partner data (Figure 2). Daily PUI’s included any patient with symptoms that raised suspicion for COVID infection, COVID exposures, and/or incomplete resolution of prior COVID infection. Based on hospital practice, movement out of the COVID-19-unit population required a not detected (ND) test result or discharge. A schematic of the model is provided in the supplemental information (Figure S2).

**Figure 2:**
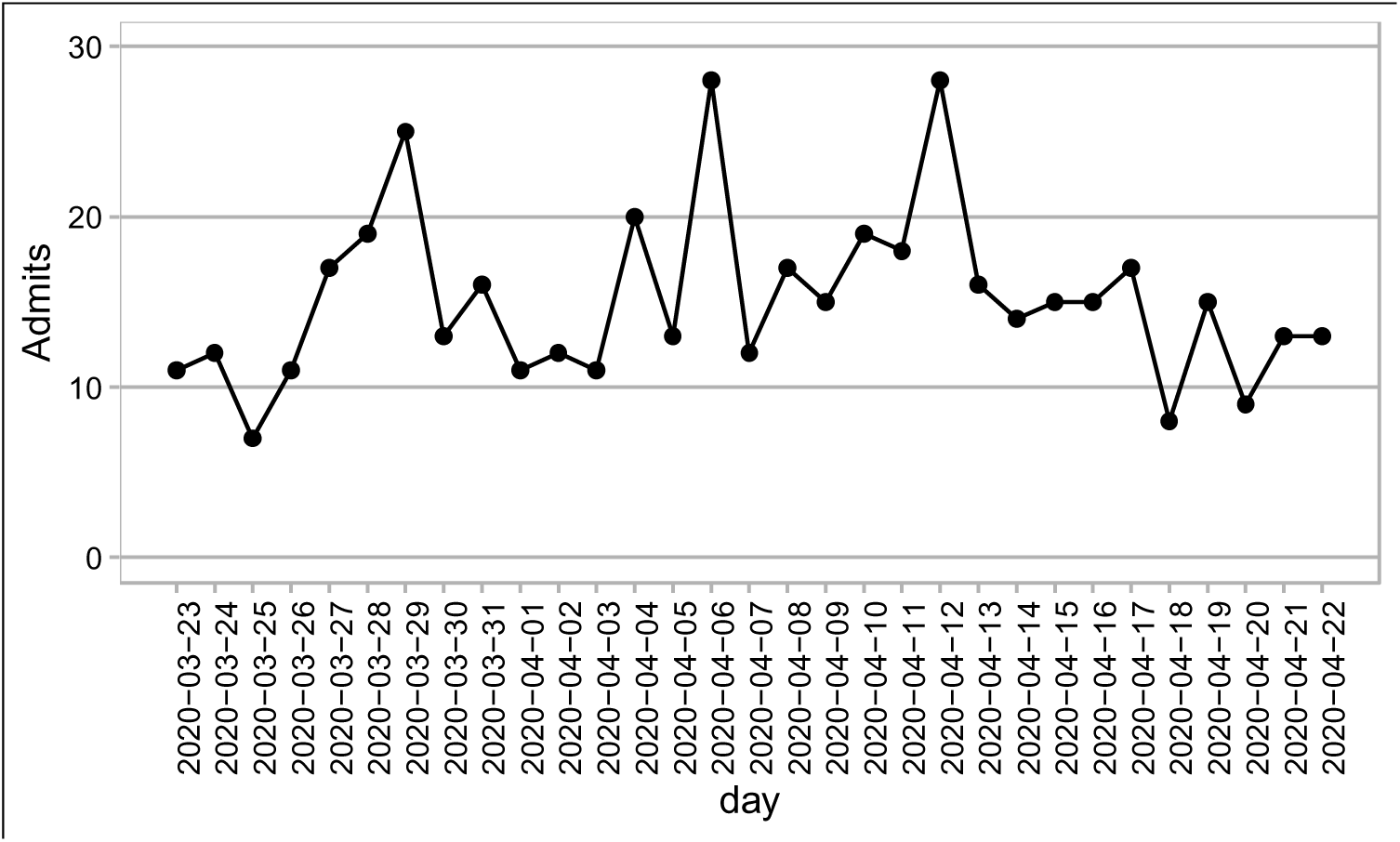
Admissions data from partner hospital shows those admitted daily for testing to the COVID-19-unit.

As our model was strictly developed to estimate potential impact on COVID-19 unit (C19U) inpatients, only the C19U population was explicitly tracked. We make the following model assumptions: 1) Specimen collection occurs on the same day as admission for testing (‘admissions’), and 2) tests are done in daily batches and returned as batches, and 3) not detected results were acted upon the same day as returned.

### COVID-19 population at timepoint *i*

The model updates the population of the COVID-19-unit daily to include daily admissions of PUI’s based on data from a hospital partner (Figure 2). Daily PUI’s included any patient with symptoms that raised suspicion for COVID infection, COVID exposures, and/or incomplete resolution of prior COVID infection. Based on hospital practice, clinicians treated these patients as potentially infected until tests results returned. Movement out of the COVID-19-unit population required a ND test result or discharge. A schematic of the model is given in supplemental Figure S2.

At each timepoint *i*, the number of individuals comprising the COVID-19-unit (C19U) population was calculated as:

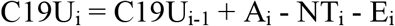

where C19U_i-1_ is the C19U yesterday (*i-1)*, A_i_ is the number of new admissions today, NT_i_ is the number of ND test results at time *i*, and E_i_ is the number of patients that egress from the COVID-19-unit at time *i*. NT_i_ was determined by patient data from hospitals based on the % positivity associated with the testing date (Supplemental Figure S3). NT_i_ was further informed by TAT so that the individuals with a ND result at time *i* are discharged at time *i+*TAT. TAT for RRTL was determined from RRTL records where the median TAT was 1.67 and the IQR was between 1.6 and 2.63, with a minimum of half a day and a maximum of just under 3 days post-collection. TAT for RRTL was thus defined daily by drawing a whole number between 1 and 3 from a uniform distribution: TAT for RRTL is given by:

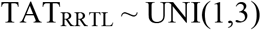

To inform the “what if RRTL had not existed” scenario for comparison, we determined the TAT of 52 patients admitted who were not tested by RRTL during a brief period when we did not operate. The TAT for these individuals was used to inform TAT in the model for non-RRTL lab entities (Figure S4). Using this data, we defined the TAT for these entities as: TAT_OTHER_ for 4% of daily C19U inpatients is 4 days, 27% of daily C19U inpatients is 5 days, 56% of daily C19U inpatients is 6 days, and 14% of daily C19U inpatients is 7 days. There was 2% (n=1 each) of patients with TAT_OTHER_ of 9 and 13 days. These were considered outliers and we capped TAT_OTHER_ at 7 days maximum. Communication from other area healthcare providers confirmed that this non-RRTL TAT was consistent with their experiences (personal communication). Accordingly, individuals admitted at day *t* would be resulted at day *(t + TAT*_*X*_) where X corresponds to either RRTL or OTHER.

Data was also provided from partner hospitals to estimate the egress from COVID-19-unit based on length of stay (LoS) data provided (Supplemental Figure S5), which includes both recovery and mortality. Patient data from sampled records of SARS-CoV-2 positive individuals (n=108) indicated that 16% of patients had a length of stay of up to 1 day, 44% had a LoS between 2 and 6 days, 17% between 7 and 10 days, 7% between 11 and 13 days, and the remaining 16% between 14 and 37 days (Supplemental Figure S5).

LoS was binned and the model parameterized according to this distribution. That is, 16% of admissions would be assigned a LoS of 1 day; 44% had a LoS defined by a randomly chosen number from a uniform distribution with a min of 2 days and a max of 6 days, etc. (details in Supplemental Table S1). Daily egress from the COVID-19-unit population was updated daily based on this distribution.

Three scenarios were considered: 1-2) RRTL capacity of only 25% or 50% of inpatient, 3) the actual RRTL capacity for testing 90% of COVID-19 inpatients and 4) no RRTL and a TAT_OTHER_ for all admissions. For all scenarios, we realized 1000 simulations and the mean number of patients in the COVID-19-unit population daily for each scenario was calculated. Projected PPE utilization impacts were calculated by estimating the number of PPE sets (e.g., gloves, masks, gowns) used daily per each individual in the COVID-19-unit as 23 per patient per day (Source: onsite counts performed by COVID-19-unit team). Cumulative savings in PPE was calculated by subtracting PPE use for scenarios 1-3 from scenario 4 to get the daily difference, which was cumulatively summed over all days of the simulation.

## Results

Longer TATs for COVID-19 tests resulted in higher C19U population numbers (Figure 3) compared to scenarios where a dedicated lab with shorter TATs. In particular, the scenario representing the RRTL workload (90% of daily COVID-19 admissions) showed a maximum one-day difference in the COVID-19-unit population of 50 patients, with 11 consecutive days of C19U occupancy having a mean of over 25 patients more compared to the null of longer TATs. Further, even smaller-scale operations had noticeable effects on the COVID-19 occupancy. When just 25% of inpatient COVID-19 tests had faster TATs, the highest one-day difference in COVID-19-unit population was 22 compared to the null, while at 50% of tests having faster TATs, the maximum impact was a 30 patient-difference compared to the null.

**Figure 3:**
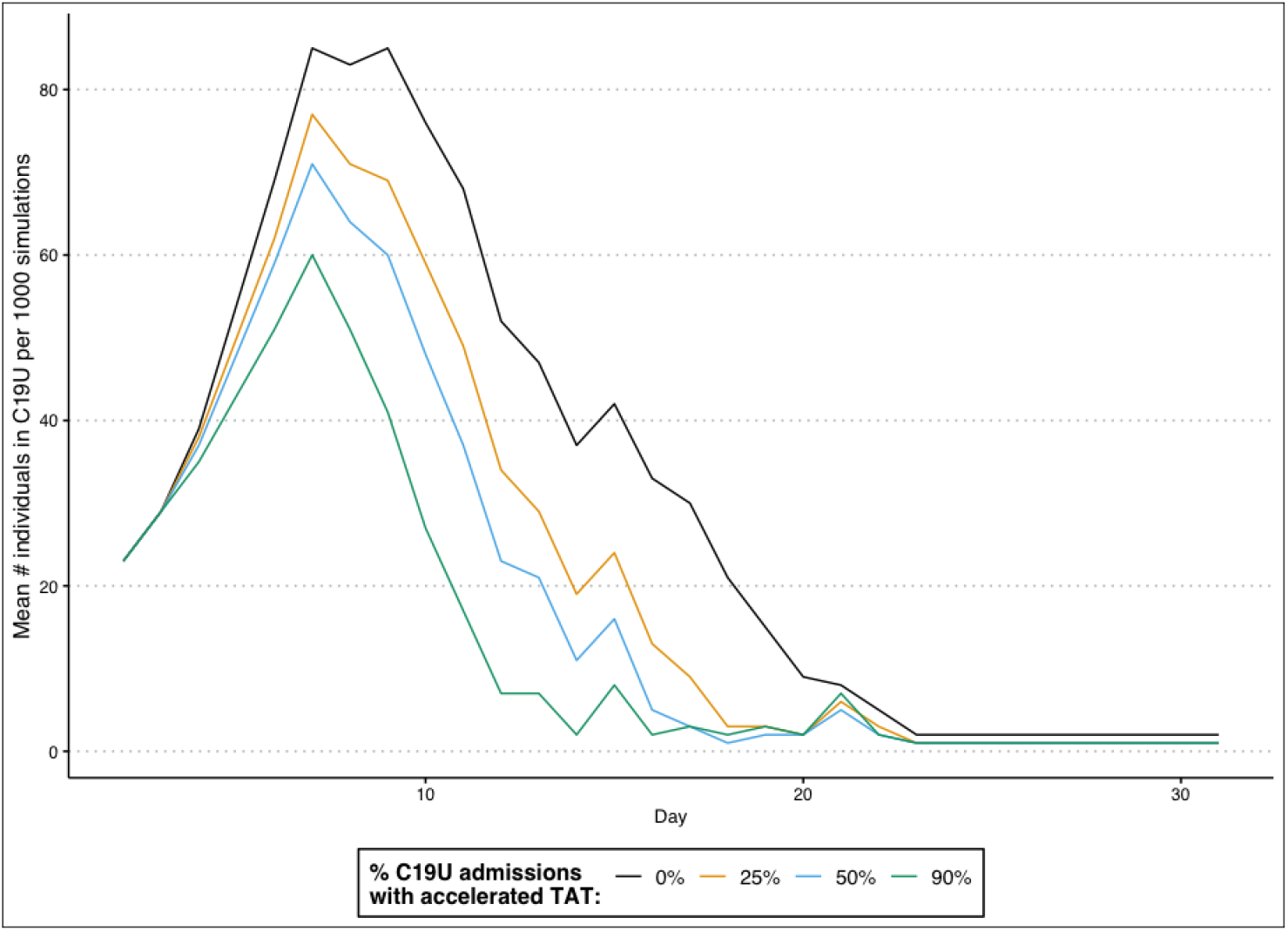
The mean daily occupancy of the C19U when an accelerated TA was available for 0% (black), 25% (yellow), 50% (blue), or 90% (green) of admissions tested.

When we calculated the impacts on PPE, at 25% faster TAT, there was a maximum savings of 506 of *each type* of PPE (masks, pairs of gloves, gowns, face shields, and goggles) at the peak day (Figure 4). Cumulatively, there was a savings of 5,681 sets of PPE (247 total patient difference). Having 50% of tests with accelerated TATs corresponded to a maximum one-day savings of 690 sets of PPE and a cumulative PPE savings of 8,096 sets (352 total patients). For the scenario most resembling RRTL, the maximum one-day savings of PPE was estimated to be 1,150 sets, with a cumulative savings of 11,316 units of PPE.

**Figure 4:**
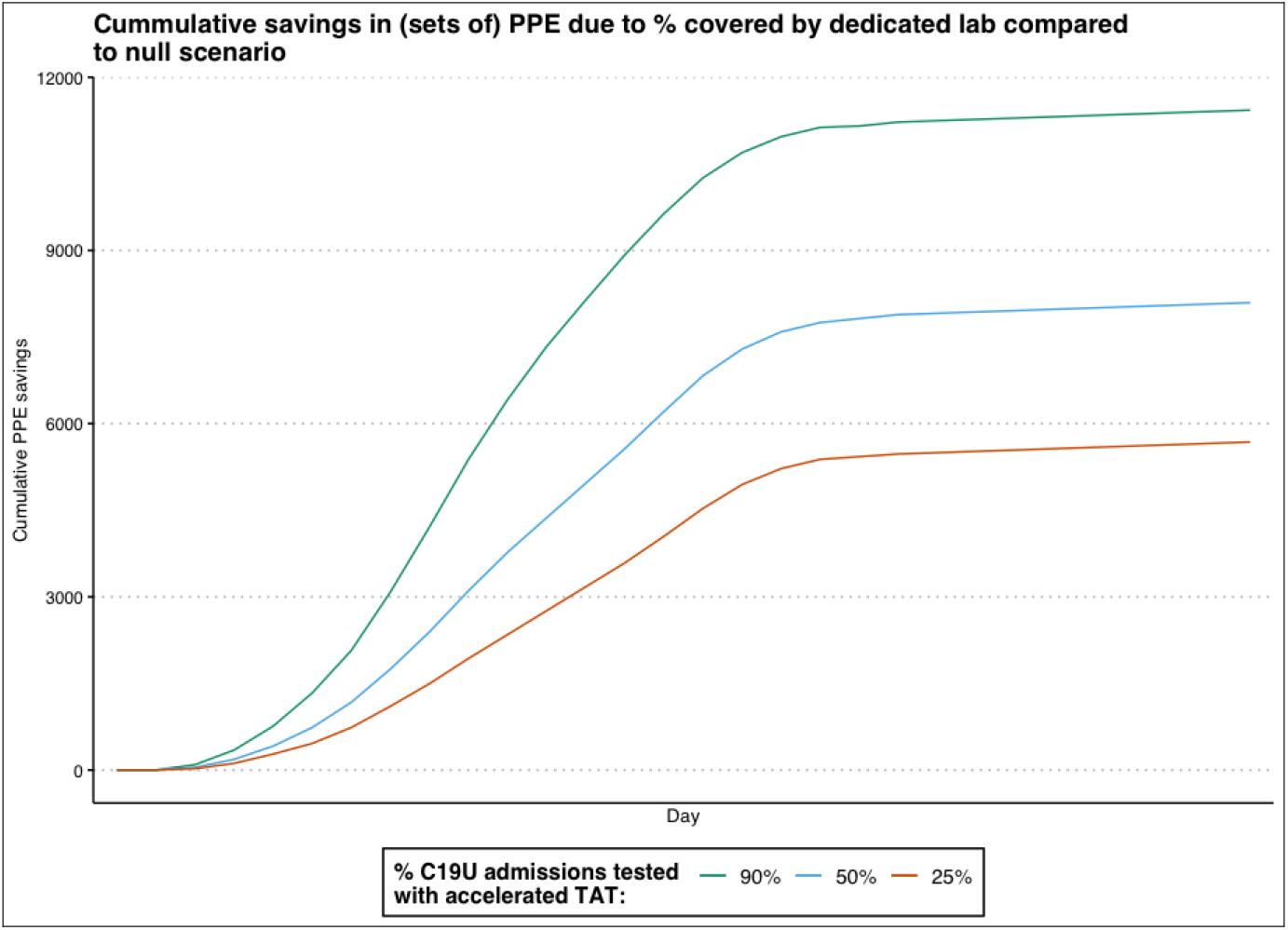
Cumulative simulated savings in PPE compared to no accelerated TAT when 25% (red line), 50% (blue line), or 90% (green line) had accelerated TATs.

## Discussion

Most state and commercial reference laboratories service very broad geographic ranges, and we have watched as demands have resulted in longer turnaround times.

The formation of RRTL as a dedicated lab allowed for focus on local demands and alleviate the burden on reference laboratories while keeping TAT short. Through the multi-sectoral partnership of academic biomedical laboratories, higher education administration, hospital physicians / physician-scientists, hospital administration, and local and state philanthropic and government officials, RRTL filled two major gaps in a crisis: availability of testing kits and CVID-19 testing capacity.

Intuitively, accelerated results would be beneficial. However, model results enable quantification of differences between what may have been had there been no RRTL laboratory. Model results suggest that faster TAT for even smaller proportions of inpatients (25%, e.g.) significantly impact hospital resource caches and bed capacity. Understanding this impact could prove critical should an expected second wave of COVID-19 illness be as big, or larger, than the first. Hospital administrators are confident that having access to the RRTL resource and rapid testing TAT significantly impacted the hospital’s ability to effectively manage COVID-19 patients, conserved critical PPE and supplies, and contributed to the overall efficiency while managing hospital operations in a pandemic. Furthermore, the impact of RRTL on the morale of local healthcare providers is immeasurable, as the impact of local expertise and personnel on the attitudes of providers proved invaluable.

Despite the positive contribution of RRTL, the repurposing of an academic virology lab for diagnostic testing – especially within the timeframe that was accomplished here – was not without challenges, including the cross-translation of regulatory lexicon and scientific communication in the setting of an academic partnership with clinical laboratory, hospital administration, finance, and operations^17^. Of the lessons learned, there are five major take-aways that other labs may encounter as COVID-19 transmission continues. First, partnerships with medical and clinical administration are critical for transition from an academic research lab to a CLIA-approved laboratory with the data and administrative capabilities to report actionable results. Proper data management and reporting architecture are necessary; integrating into existing frameworks used by hospitals was the most efficient route. Second, careful consideration should be made about converting what is often older laboratory space into a lab designed to have a clean-to-dirty flow (one-way flow) ^18^. The RRTL process involved several discrete checkpoints to keep contamination from occurring, including daily decontamination of all laboratory spaces, and development of process flow that created as close to a one-way flow as possible. Third, Louisiana’s outbreak occurred early relative to most states, though it was somewhat coincident with the large-scale outbreak in NYC and the ongoing transmission in Washington state ^4,19^. Even so, we experienced significant supply chain issues despite early preparation prior to the case surge in both Louisiana and NYC, that required direct resolution of issues with the vendors themselves. We strongly suggest that long-term supply chain continuity be a major part of planning and regularly updated, as expectation is that a second wave could cause interruptions and/or delays.

Fourth, reasonable relaxation of regulatory requirements was a major determinant in the speed at which RRTL came online. While regulatory relief was available midway through the process, RRTL could have come online days sooner had the regulations relaxed sooner. We suggest that as a matter of federal and state policy during times of national emergencies, academic labs that are willing and able should be enabled for fast-tracked approvals under the guidance of professional clinical laboratory partners. This might require existing agreements between willing laboratories and local hospital laboratory directors and/or pathologists to ensure quality control and adherence to best practices. Having in place an existing fast track for qualified academic labs to transition will allow these collaborative efforts to come online quickly to meet the needs of the community. Lastly, the efforts described herein were made possible only by the forethought of the physician scientist to investigate capabilities at LSU, leading to the formation of RRTL. Local hospitals should follow such lead with the understanding that academic laboratories have unique expertise that may be useful in public health emergencies. Hospital administrators and physicians should engage nearby academic laboratories and work on formulating collaborations and agreements in preparation for future pandemics.

In summary, the formation of a targeted testing lab focused primarily on COVID-19 patients with faster-than-average TAT was critical for maintaining control over hospital capacity and resources. Given this dedicated system, RRTL was able to alleviate stress on state and hospital laboratories for the region serviced and prevent unnecessary stress on healthcare associated with testing backlogs. Academic biomedical laboratories are resourced with the capabilities to have significant impacts for their communities during public health emergencies. Similar to the reserve forces of the United States Armed Forces and National Guard, public health emergency responses should be enabled to draw upon a reserve of healthcare and biomedical research professionals embedded within communities who have the expertise, experience, and knowledge to fill critical gaps in capacity when necessary.

## Data Availability

All relevant data is contained within the manuscript and supplementary information file.

## Acknowledgements

We would like to thank the following individuals for their support and insights into this effort from lab stand-up to manuscript development: Governor John Bel Edwards, Mayor Sharon Weston Broom, John Spain, Dr. Sam Bentley, Dr. Stacia Haynie, Dr. Tom Galligan, Dr. George Karam, Dr. Joel Silverberg, Dr. Elizabeth Floyd, Jim Teague, Thiago Menegon, Senator Bill Cassidy, M.D., Senator John Kennedy, Walter Braud, Amy Landry, Dr. Christopher Mores, Ginger Gutner, Evelyn R. Christofferson, Christine LeBeouf, Deekshith Mandala, Milad Amini, Michael Zarruk, LeaAnn Teague, all those who took care of our families while we were thoroughly occupied in this endeavor, and all the cats who have zoom-bombed the world over.

## References

1. Coronaviridae Study Group of the International Committee on Taxonomy of V. The species Severe acute respiratory syndrome-related coronavirus: classifying 2019-nCoV and naming it SARS-CoV-2. Nat Microbiol 2020; 5(4): 536–44.

2. Weiss SR, Navas-Martin S. Coronavirus pathogenesis and the emerging pathogen severe acute respiratory syndrome coronavirus. Microbiol Mol Biol Rev 2005; 69(4): 635–64.

3. Su S, Wong G, Shi W, et al. Epidemiology, Genetic Recombination, and Pathogenesis of Coronaviruses. Trends Microbiol 2016; 24(6): 490–502.

4. Holshue ML, DeBolt C, Lindquist S, et al. First Case of 2019 Novel Coronavirus in the United States. N Engl J Med 2020; 382(10): 929–36.

5. Paranthaman K, Conlon CP, Parker C, McCarthy N. Resource allocation during an influenza pandemic. Emerg Infect Dis 2008; 14(3): 520–2.

6. Sun Ea. Multi-objective optimization models for patient allocation during a pandemic influenza outbreak. Elsevier 2014; 51:350–9.

7. Paolo Pasquariello SS. Excess Mortality from COVID-19: Lessons Learned from the Italian Experience. PrePrint 2020.

8. Boccia S, Ricciardi W, Ioannidis JPA. What Other Countries Can Learn From Italy During the COVID-19 Pandemic. JAMA Intern Med 2020.

9. Mirco Nacoti ea. At the Epicenter of the COVID-19 Pandemic and Humanitarian Crises in Italy: Changing Perspectives. on Preparation and Mitigation. N Engl J Med Catalyst 2020.

10. Qiu H, Tong Z, Ma P, et al. Intensive care during the coronavirus epidemic. Intensive Care Med 2020; 46(4): 576–8.

11. Bnaya Gross ea. Spatio-temporal propagation of COVID-19 pandemics. MedRxiv 2020.

12. Policy for Diagnostic Tests for Coronaivurs Disease-2019 during the Public Health Emergency. In: Administration USFD, editor.; 2020.

13. Brangham W. The supply chain fiasco that has derailed COVID-19 testing in the U.S. In: Woodruff J, editor. PBS News Hour; 2020.

14. Anderson M, Pfeiffer S, Van Woerkom B. Despite Early Warnings, U.S. Took Months To Expand Swab Production For COVID-19 Test. 2020. https://www.npr.org/2020/05/12/853930147/despite-early-warnings-u-s-took-months-to-expand-swab-production-for-covid-19-te (accessed May 30, 2020 2020).

15. Harris PA, Taylor R, Thielke R, Payne J, Gonzalez N, Conde JG. Research electronic data capture (REDCap)--a metadata-driven methodology and workflow process for providing translational research informatics support. J Biomed Inform 2009; 42(2): 377–81.

16. Harris PA, Taylor R, Minor BL, et al. The REDCap consortium: Building an international community of software platform partners. J Biomed Inform 2019; 95:p 103208.

17. Lefferts JA, Gutmann EJ, Martin IW, Wells WA, Tsongalis GJ. Implementation of an Emergency Use Authorization Test During an Impending National Crisis. J Mol Diagn 2020; 22(7): 844–6.

18. Organization WH. Dos and Don’ts for molecular testing. 2018. https://www.who.int/malaria/areas/diagnosis/molecular-testing-dos-donts/en/ (accessed May 17, 2020 2020).

19. Flores S, Gavin N, Romney ML, et al. COVID-19: New York City pandemic notes from the first 30 days. Am J Emerg Med 2020.

