## Supplemental Information for "Reduced turnaround times through multi-sectoral collaboration during the first surge of SARS-CoV-2 in Louisiana, March-April 2020"

Multisectoral collaboration for pandemic response and operational support of critical care and emergency departments: Supplemental Information

**Supplemental Figure S1:** Typical daily schedule for RRTL resulting in decreased turnaround times for COVID-19 inpatients at area hospitals.

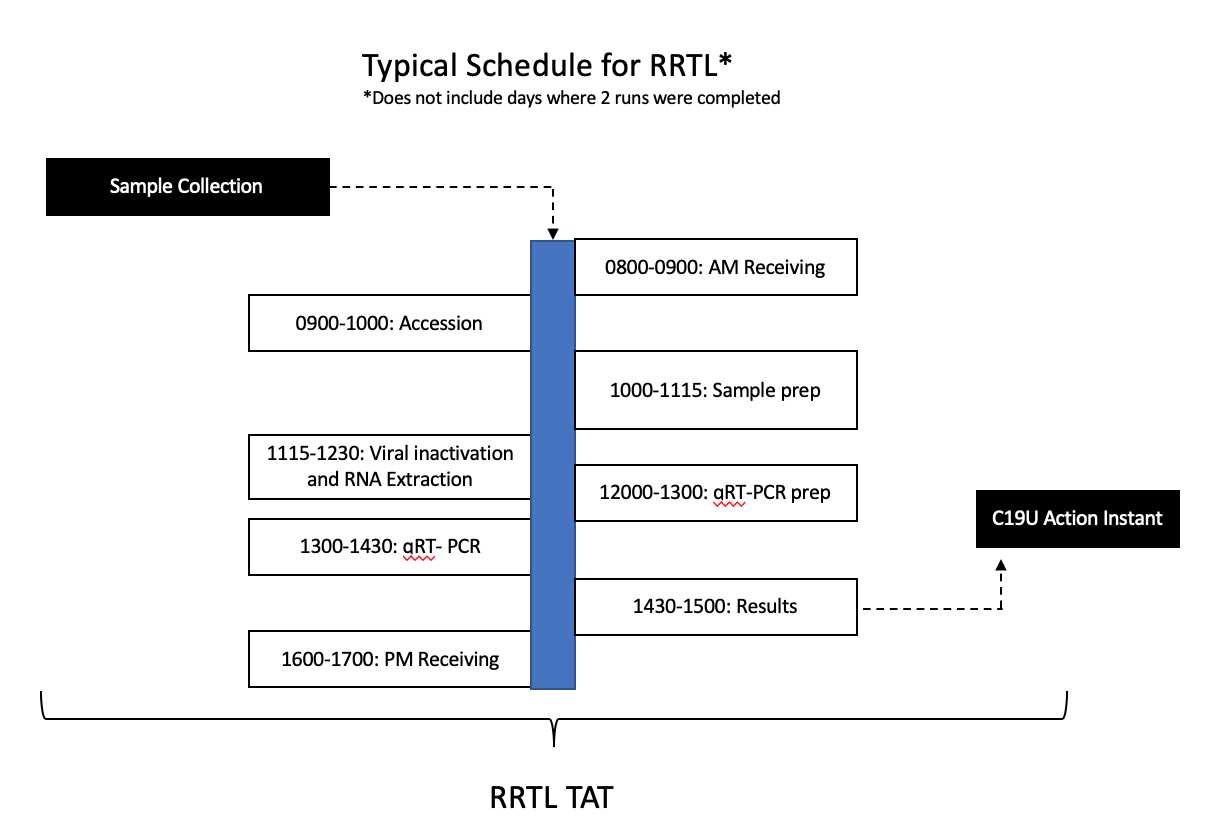

**Supplemental Figure S2: Model Schematic**

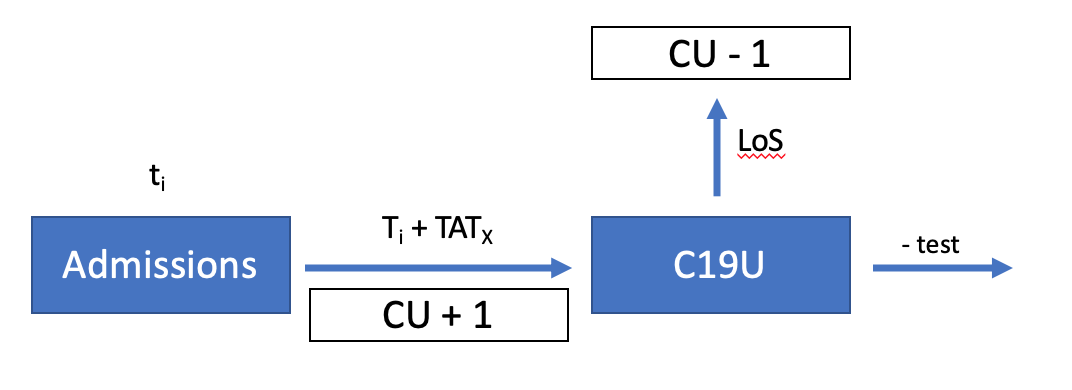

**Supplemental Figure S3:** Percent C19U positive individuals according to admitted for testing date.

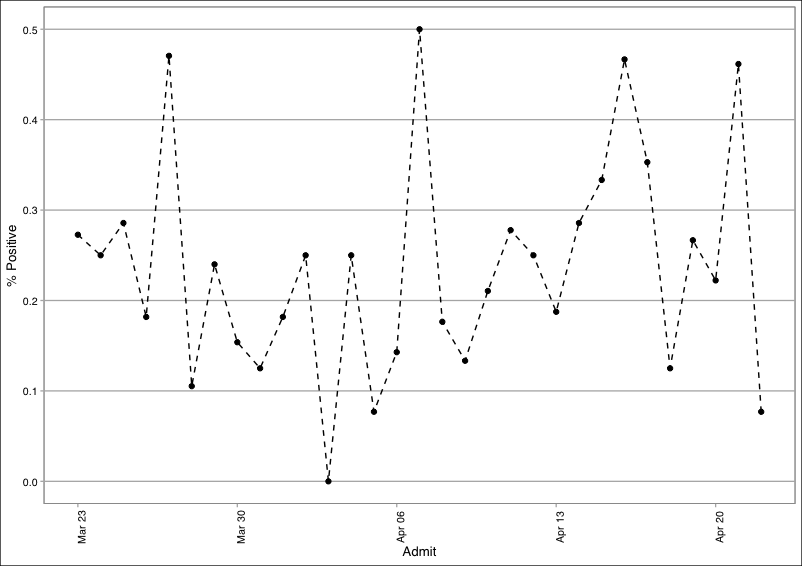

**Figure S4**: Turnaround time for non-RRTL tests during the RRTL offline period. There were two tests in each of the 9 and 13 day TAT, but these were considered outliers and only TAT between 4 and 7 days were used.

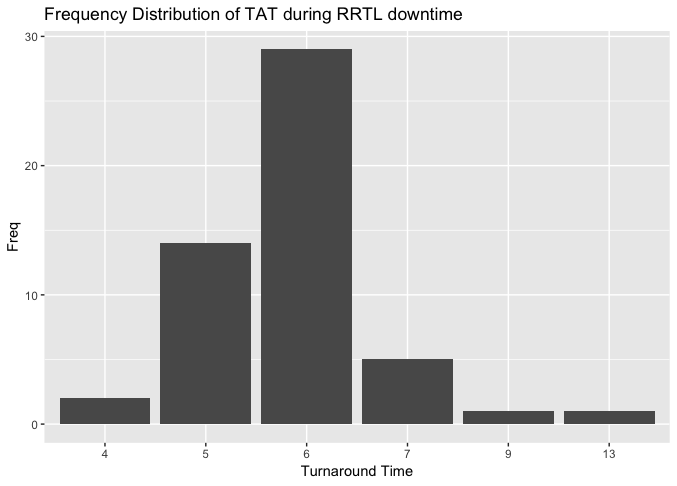

**Supplemental Figure S5**: A) Box plot showing the distribution of length of stay binned with the proportion of patients on the y-axis. B) Individual percentages of LoS from the patients in our data and C) Boxplot with the IQR of [2,10].

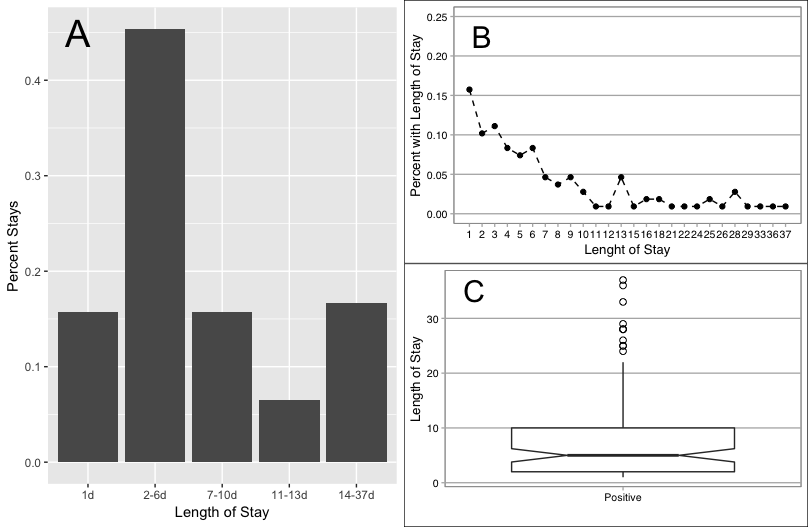

**Table S1**: Length of stays from 108 sampled inpatients determined by record examination and binned into categories based on similarity of proportions.

| Length of stay (days) | proportion | bin |
| --- | --- | --- |
| 1 | 0.16 | 1 |
| 2 | 0.10 | 2 |
| 3 | 0.11 | 2 |
| 4 | 0.08 | 2 |
| 5 | 0.07 | 2 |
| 6 | 0.08 | 2 |
| 7 | 0.05 | 3 |
| 8 | 0.04 | 3 |
| 9 | 0.05 | 3 |
| 10 | 0.03 | 3 |
| 11 | 0.01 | 4 |
| 12 | 0.01 | 4 |
| 13 | 0.05 | 4 |
| 15 | 0.01 | 5 |
| 16 | 0.02 | 5 |
| 18 | 0.02 | 5 |
| 21 | 0.01 | 5 |
| 22 | 0.01 | 5 |
| 24 | 0.01 | 5 |
| 25 | 0.02 | 5 |
| 26 | 0.01 | 5 |
| 28 | 0.03 | 5 |
| 29 | 0.01 | 5 |
| 33 | 0.01 | 5 |
| 36 | 0.01 | 5 |
| 37 | 0.01 | 5 |
